# Statistical analysis of three data sources for Covid-19 monitoring in Rhineland-Palatinate, Germany

**DOI:** 10.1101/2023.09.21.23295894

**Authors:** Maximilian Pilz, Karl-Heinz Küfer, Jan Mohring, Johanna Münch, Jarosław Wlazło, Neele Leithäuser

**Author notes:** Correspondence: Maximilian Pilz <.de>.

## Abstract

In Rhineland-Palatinate, Germany, a system of three data sources has been established to track the Covid-19 pandemic. These sources are the number of Covid-19-related hospitalizations, the Covid-19 genecopies in wastewater, and the prevalence derived from a cohort study. This paper presents an extensive comparison of these parameters. It is investigated whether wastewater data and a cohort study can be valid surrogate parameters for the number of hospitalizations and thus serve as predictors for coming Covid-19 waves. We observe that this is possible in general for the cohort study prevalence, while the wastewater data suffer from a too large variability to make quantitative predictions by a purely data-driven approach. However, the wastewater data as well as the cohort study prevalence are able to detect hospitalizations waves in a qualitative manner. Furthermore, a detailed comparison of different normalization techniques of wastewater data is provided.

## 1 Introduction

Wastewater surveillance is an important tool to investigate the current status of the Covid-19 pandemic (Feng et al. 2021) and has the potential to be used for further viruses as well (Diamond et al. 2022). The course of the Covid-19 pandemic is investigated in many countries by wastewater surveillance. Among others, USA (Duvallet et al. 2022), New Zealand (Hewitt et al. 2022), the Netherlands (Langeveld et al. 2023) and Switzerland (Cariti et al. 2022) used this tool as a means of monitoring the detected genecopies in their sewage systems. Among the intended advantages of wastewater monitoring are its independence of individual testing which varies with political guidelines, its moderate costs, its anonymous character, and that there is no additional effort for the population. However, there are various sources of variation that impact the amount of measured genecopies (Wade et al. 2022).

A high viral load does not necessarily mean a major public health burden. The really critical quantity is the number of persons who have to go to hospital because of the virus. However, this quantity is no longer recorded as Covid-19 tests in hospitals have mainly been stopped (Robert Koch-Institut 2023) and the pandemic will be more and more monitored via wastewater. Before, the rate of infected, hospitalized patients was considered a very important measure of the burden of disease in the population. Furthermore, hospitals were regarded as a source of infection and regular testing of all employees and patients was considered as a means of containment. Since these information will not be available in the future, one major goal of this publication is to investigate whether there is a relationship between viral load and hospitalization from the period where both measurements were available.

Besides wastewater surveillance, the German federal state Rhineland-Palatinate established a cohort study in which voluntary citizens regularly tested themselves on Covid-19 and reported the results to the clinical trial organization team. Contrary to the officially reported case data, which is biased by testing resources and motivation, this study data is much more representative and bypasses the problem of a large number of undetected cases.

In this paper, we compare the results of the data sources wastewater surveillance, cohort study, and the reported number of Covid-19-related hospitalizations. All three sources are available in Rhineland-Palatinate between January and April 2023. We illustrate the effect of the normalization technique in wastewater and investigate whether a data-based prediction from wastewater surveillance could be an early-warning system for the detection of pandemic waves. While there are many publications that compare wastewater data to officially reported test data, to the best of our knowledge, to date, there are no comparative results from wastewater and large population cohorts.

In Section 2, we present the three data sources in more detail and describe the applied statistical methodology. In Section 3, the results are presented. We conclude with a discussion in Section 4.

## 2 Methods

### 2.1 Data sources

#### 2.1.1 Wastewater

Wastewater was taken twice per week in 15 sewage plants in Rhineland-Palatinate by taking 24-hours composite samples, following the same guidelines as were developed for the EU project ESI-CorA (Robert Koch Institute 2023). For each measurement, the number of N1 and N2 genecopies, respectively, was determined. Those are the two gene targets of the SARS-CoV-2 virus that are commonly used in laboratory testing for Covid-19 (Feng et al. 2021). Besides this, a reference virus, the Pepper mild mottle virus (PMMoV), was measured. This is a plant RNA virus that is known to be a good reference in wastewater to be compared with other viruses (Kitajima, Sassi, and Torrey 2018). The laboratory followed the manufacturer’s standard protocol for the Promega Maxwell (R) RSC Enviro Total Nucleic Acid Kit for extracting the viral information.

Since wastewater occurrence of viruses may vary with the rainfall, the water volume of each measurement day at each sewage plant was measured as well. In addition, some wastewater related parameters were collected. Those are the water temperature, the pH value in water, the chemical oxygen demand, the water conductivity, and total organic carbon. The respective sewage plants together with the number of connected inhabitants and the plants’ dry-weather flows are available as well. To investigate possible weather-related influences, we added information on the air temperature and the precipitation that is available online by the German Meteorological Service (Service 2023). Since these parameters can not be aggregated on federal state level, they are only used in Section 3.4.

While the wastewater surveillance in Rhineland-Palatinate is still ongoing, we use its measurement values between December 2022 and April 2023, thus for 5 months, corresponding to the availability of the other data sources. We have this data for each treatment facility individually. A publicly available extract of this data is available online (Rheinland-Pfalz 2023). The participating sites include the five cities that were selected for the cohort study.

#### 2.1.2 Cohort study

In 2022 and 2023, the University Medical Center of the Johannes Gutenberg-University Mainz conducted a cohort study (“SentiSurv”) on behalf of the Ministry of Science and Health of Rhineland-Palatinate (Wild 2023b). A cohort was drawn randomly from the five largest cities in Rhineland-Palatinate, namely Mainz, Ludwigshafen, Koblenz, Trier, and Kaiserslautern. These cities are uniformly distributed across the federal state. The participants are almost representative in terms of gender and age except for children who have been excluded for legal reasons. During the course of the first months, all five cohorts reached their targeted size of 2800 volunteers each. In addition to the regular completion of questionnaires, participants have performed a rapid test on two fixed days each week and have reported the outcome in a mobile app. Starting from January 2023, the number of participants included in the study was large enough to allow a statistically sound interpretation.

The results of the cohort study are available between January 2023 and April 2023, thus for 4 months. We have this data for each of the tested cities individually. The data has been displayed online during the course of the study (Wild 2023a).

#### 2.1.3 Hospitalizations

As an official reference value, the number of Covid-19 related hospitalizations are used. Those are the number of hospitalized persons that were tested positive on Covid-19 for a given day. Note that this differs from the commonly reported number of newly admissioned patients with a positive test. A Covid-19 test was mandatory in hospitals in Rhineland-Palatinate until March 31, 2023, and therefore, the hospitalizations are available between December 2022 and March 2023, thus for 4 months. The data was collected on a weekdaily basis from the Ministerium für Wissenschaft und Gesundheit (Ministry of Science and Health) in Rhineland-Palatinate by calling the individual hospitals and ask for their positively tested patients. We only have these values for the whole of Rhineland-Palatinate, not broken down to cities. To the best of our knowledge, the numbers are not officially reported.

### 2.2 Data analysis

#### 2.2.1 Parameter computation

From the collected parameters, some additional values could be derived. To appropriately depict the variation in N1 and N2 genecopies simultaneously, one may report their mean instead of only one of these values. Furthermore, for all three genecopies (N1, N2, and their mean), one can report their absolute value, their value per ml (i.e. normalized by the flow volume adjusted by the sewage plant’s dry-weather flow), and their value in relation to the reference virus PMMoV. The latter value is computed as genecopies */* PMMoV *·* 100,000. Three genecopies and three normalization techniques result in a number of nine parameters that can be computed for the wastewater.

The nine numbers defined above (N1, N2, and mean, reported as absolute value, per ml, and per PMMoV, respectively) have to be normalized with the number of inhabitants in order to combine the values per sewage plant to summary values for the whole of Rhineland-Palatinate. For the three genecopies (N1, N2, and mean), this normalization happens in the same manner. Therefore, in the following formulas, the normalization is presented for the absolute value of genecopies, for the genecopies per ml, and the genecopies per PMMoV, where genecopies stands for N1, N2, or their mean. Since the total number of genecopies is an absolute value, one can compute

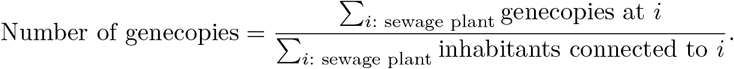

For the relative parameters, the numerator in the above formula would not necessarily increase with increasing number of inhabitants. This implies that sewage plants with small population may be over-represented. Therefore, for these parameters, the following formulas hold:

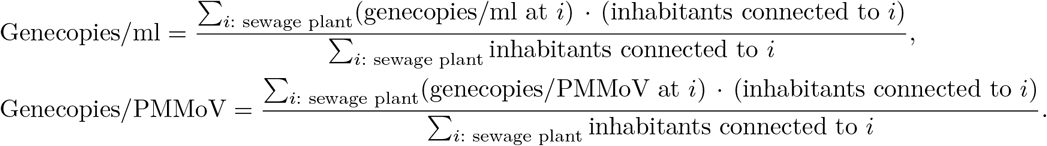

The resulting values are multiplied by 100,000 in order to report the values per 100,000 inhabitants.

For the cohort study, the daily prevalence is reported. This means that the number of positively tested participants is divided by the total number of participants. The resulting value is multiplied by 100,000 to obtain the prevalence per 100,000 inhabitants. The cohort study Covid tests were done on Sunday on Wednesday. We denote the Sunday measurement as first measurement and the Wednesday measurement as second measurement of a week.

In order to match the hospitalization data to the two measurements per week, the mean of the number of hospitalizations of Monday, and Tuesday for week measurement one and of Wednesday, Thursday, and Friday for week measurement two are computed, respectively.

#### 2.2.2 Statistics

The resulting parameters can be depicted as point plots. To measure the uncertainty in the data, confidence intervals for all values are needed. The prevalence can be interpreted as a rate and, therefore, Wilson confidence intervals for rates (Wilson 1927) can be computed and multiplied by 100,000 as well. For continuous parameters, we compute the confidence interval for the mean by assuming a normal distribution. To this end, the standard deviation of the measurements has to be estimated. We do this applying a time-shifted concept by computing for each measurement the standard deviation of the respective measurement, and the two measurements before and after this measurement. For the hospitalizations, multiple values are summarized to compute one value per measurement (cf. Section 2.2.1). These values are used to compute the standard deviation for each measurement. Since the reported values cannot be negative, the lower bounds of the confidence intervals are cut at zero.

To illustrate time trends, a smoothing method is needed. We applied locally estimated scatterplot smoothing (LOESS) regression. This method fits for each data point a linear regression in a pre-defined neighborhood of this point and predicts the point by this linear regression. By this neighborhood approach, a new linear model is fitted for each data point and thus a local smoothing is performed. The proportion of data points from the entire dataset that is used for the local linear regression is called span. In this paper, we used a span of 50% for the wastewater data and 60% for the comparison of all data due to the lower number of datapoints for the cohort study and hospitalization.

In order to investigate if there is a time shift between different parameters, time lag correlations are computed between the raw values of these parameters. This means that one of the two parameters to be compared is shifted by a time step of *l* and the correlation between the two resulting time series is computed. The result is the correlation with time lag *l*. This approach will be used to compare if there is a time shift between prevalence, hospitalizations, and wastewater data.

We tried to use the available data to build prediction models in a data-driven manner, i.e. without the knowledge of any biological background. First, we investigated whether one can predict the number of hospitalizations by wastewater data and the cohort study prevalence. Second, we analyzed the predictive capability of the wastewater data to predict the cohort study prevalence. To this end, regression models were applied. We fitted random forests (Breiman 2001) to predict the outcome of interest. The number of trees in each forest was set to 5000 and the candidates at each split were set to 5. In order to avoid overfitting, leave-one-out cross-validation was applied. This means that for each measurement, a random forest was fitted on all other data points, but not this measurement, and the measurement was then predicted by the resulting random forest.

To make the scenario realistic, past values of the variables were included as covariates into the regression models. Those past values are denoted with lag *i* when talking about the timepoint *i* measurements before the current observation. Feature importance was computed by a random forest fit on the full datasets to investigate which covariates influence the prediction remarkably. This feature importance was calculated as the factor by which the random forests prediction error increases when the respective feature is shuffled within the underlying dataset. As error measure, the robust mean squared error (RMSE) was used.

Data was analyzed and visualized using the statistical software R (R Core Team 2023), version 4.3.0, and the tidyverse packages (Wickham et al. 2019). Random forests were fit with the R package randomForest (Liaw and Wiener 2002) and the R package iml (Molnar, Bischl, and Casalicchio 2018) was applied for the calculation of feature importance.

## 3 Results

We present our results as four main findings that are given as the respective subchapter headings.

### 3.1 Finding 1: Trends in wastewater are present irrespective of genecopy type and normalization technique

Figure 1 shows the three genecopies (N1, N2, and their mean) together with three normalization techniques (absolute value, per ml, and per PMMoV).

**Figure 1.**
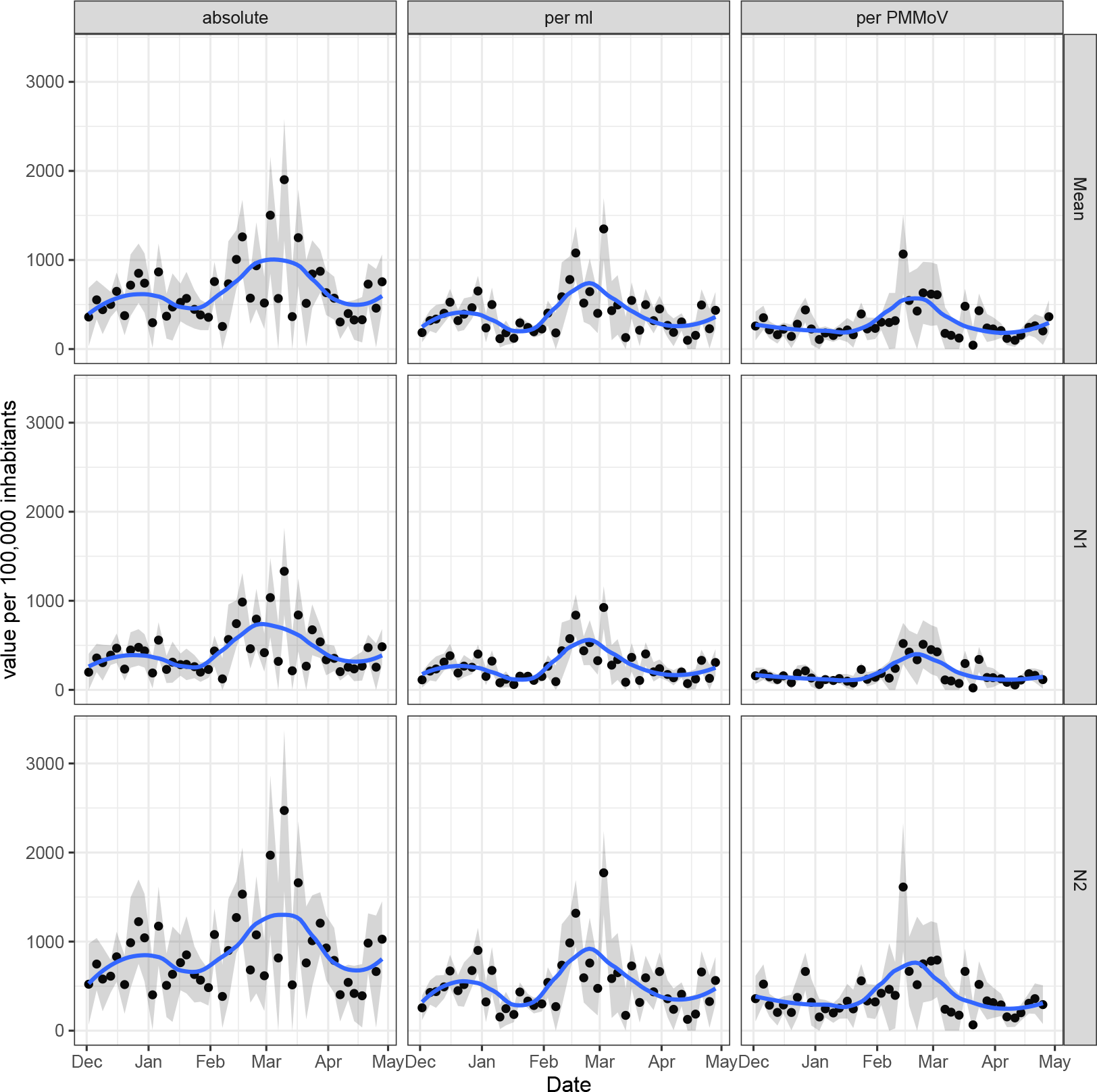
Wastewater values (mean, N1, and N2) in dependence of normalization technique. Main trends are detectable in all types of genecopies and all normalization techniques. Normalizing by flow or PMMoV slightly regularizes the curve.

Regarding trends, all nine panels show the same pattern with a small wave in late 2022 and a second, larger, wave starting in February 2023. The N2 values are larger than the N1 values but show the same pattern. Normalizing the data by the flow or the reference virus compresses the data without changing the depicted trends. Therefore, it seems to act as regularization. When normalizing the data by the flow or the PMMoV, the second wave reaches its peak before March 2023 while the absolute virus value continues increasing until mid March 2023. This behavior is observed for N1, N2, and their mean analogously. After the peak, all curves decrease and show a moderate increase in the last measurements.

### 3.2 Finding 2: Qualitative trends are observed in all three data sources

Figure 2 compares the three data sources genecopies (normalized per ml), prevalence from the cohort study, and number of hospitalizations according to their availability described in Section 2.1.

**Figure 2.**
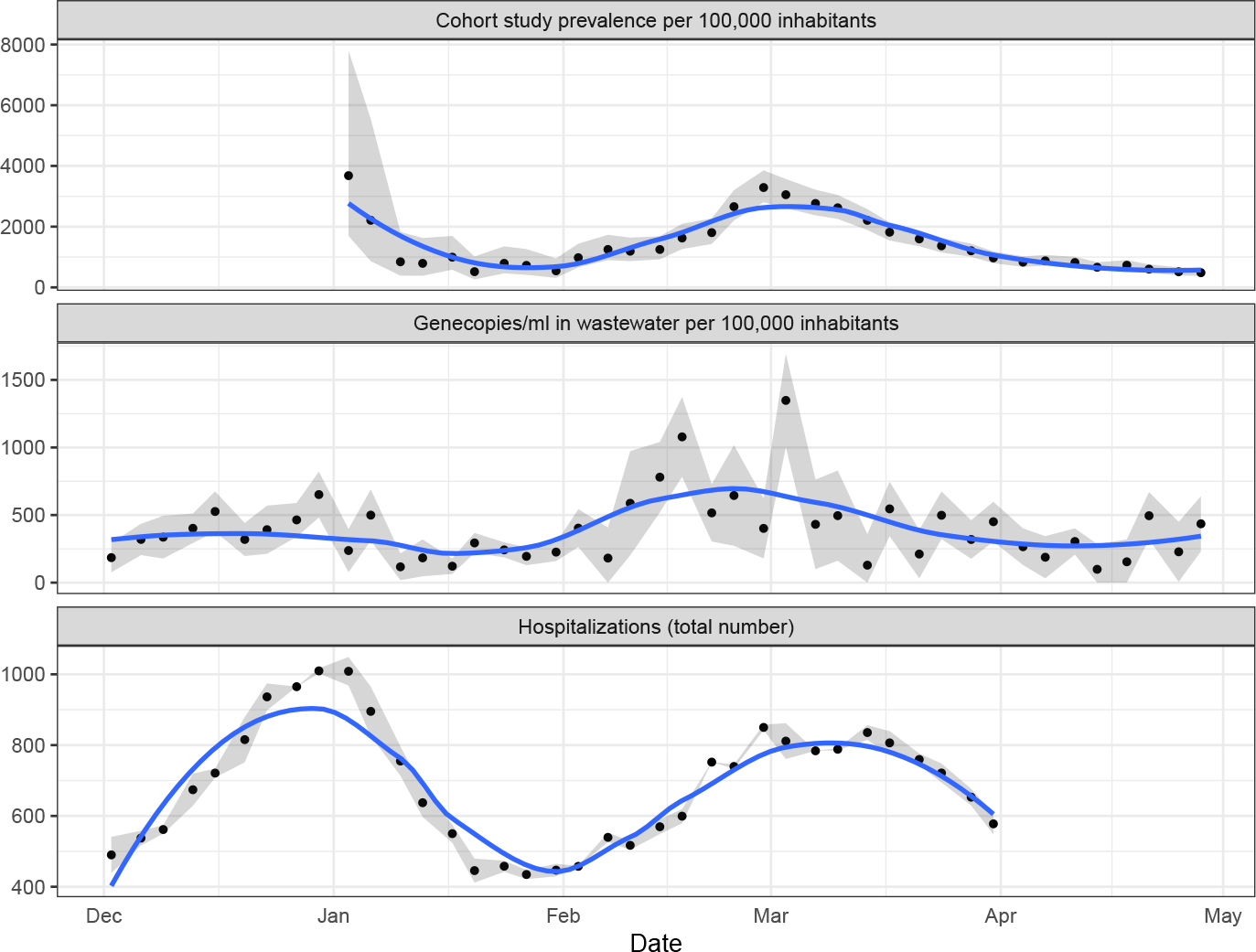
Comparison of prevalence, genecopies, and hospitalizations. The main wave is observable in all three parameters.

There are two hospitalization waves. The first, slightly higher wave, has its peak around New Year. After a minimum in late January, the second wave has its peak in the first half of March. Due to missing data from 2022, the cohort study prevalence only captures the second wave. The peak of this wave is reached about 7 days earlier than in the hospitalizations. This indicates that the cohort study may be a good tool to detect hospitalization waves with some time lead. The wastewater data capture both waves but in different intensity. The second peak is here distinctly higher than the second one. Of note, the wastewater values show a much larger variance than the other two data sources. This complicates the interpretation of single values and the early detection of potential virus waves.

### 3.3 Finding 3: Cohort study prevalence and wastewater data allow prediction of hospitalizations

Figure 2 indicates that hospitalization waves may be announced earlier in the wastewater and the cohort study prevalence, respectively. Table 1 shows the time lag correlation between the raw values of the three data sources by reporting two values for the wastewater data analogously to Figure 2. The time lag is defined such that the first mentioned value is shifted to the left by the respective time lag.

**Table 1:** Time lag correlation between the data sources. Time lag is given in measurements.

| Time lag | Gen.ml/Hosp. | Pre./Hosp. | Gen.ml/Pre. |
| --- | --- | --- | --- |
| -4 | 0.382 | 0.292 | 0.468 |
| -3 | 0.481 | 0.441 | 0.591 |
| -2 | 0.412 | 0.572 | 0.613 |
| -1 | 0.370 | 0.704 | 0.581 |
| 0 | 0.219 | 0.765 | 0.445 |
| 1 | 0.140 | 0.629 | 0.419 |
| 2 | -0.079 | 0.415 | 0.177 |
| 3 | -0.192 | 0.198 | -0.005 |
| 4 | -0.397 | -0.046 | -0.125 |

The genecopies per ml show a slight correlation with the number of hospitalizations 14 to 3 days (4 to 1 measurements) later. The highest correlation values are observed between the cohort study prevalence and the hospitalizations within a time difference of 0 to 7 days (0 to 1 measurements). Genecopies per ml and the prevalence are positively correlated as well where the prevalence seems to fit better to the hospitalizations than the genecopies per ml.

Eventually, we investigated whether the number of hospitalizations can be predicted by the cohort study prevalence, raw values of the genecopies, and the genecopies per ml including the past three values of these parameters. As described in Section 2.2.2, random forests with leave-one-out cross-validation were fitted.

Figure 3 shows the results of the hospitalization prediction. In the left panel, the true numbers of hospitalizations are plotted together with the random-forest-predicted hospitalizations. The prediction is able to detect the wave in the first weeks of March, including the increase before and the decrease after the peak. The predicted values are slightly tightened compared with the true values. The right panel shows the feature importances. Clearly, the three most important parameters are the cohort study prevalences zero to two measurements (thus 0 to 10 days) before the current measurement. This corresponds with the findings of Figure 2 and Table 1 that there is a certain time shift between the prevalence and the number of hospitalizations. The genecopies in wastewater do not influence the outcome remarkably.

**Figure 3.**
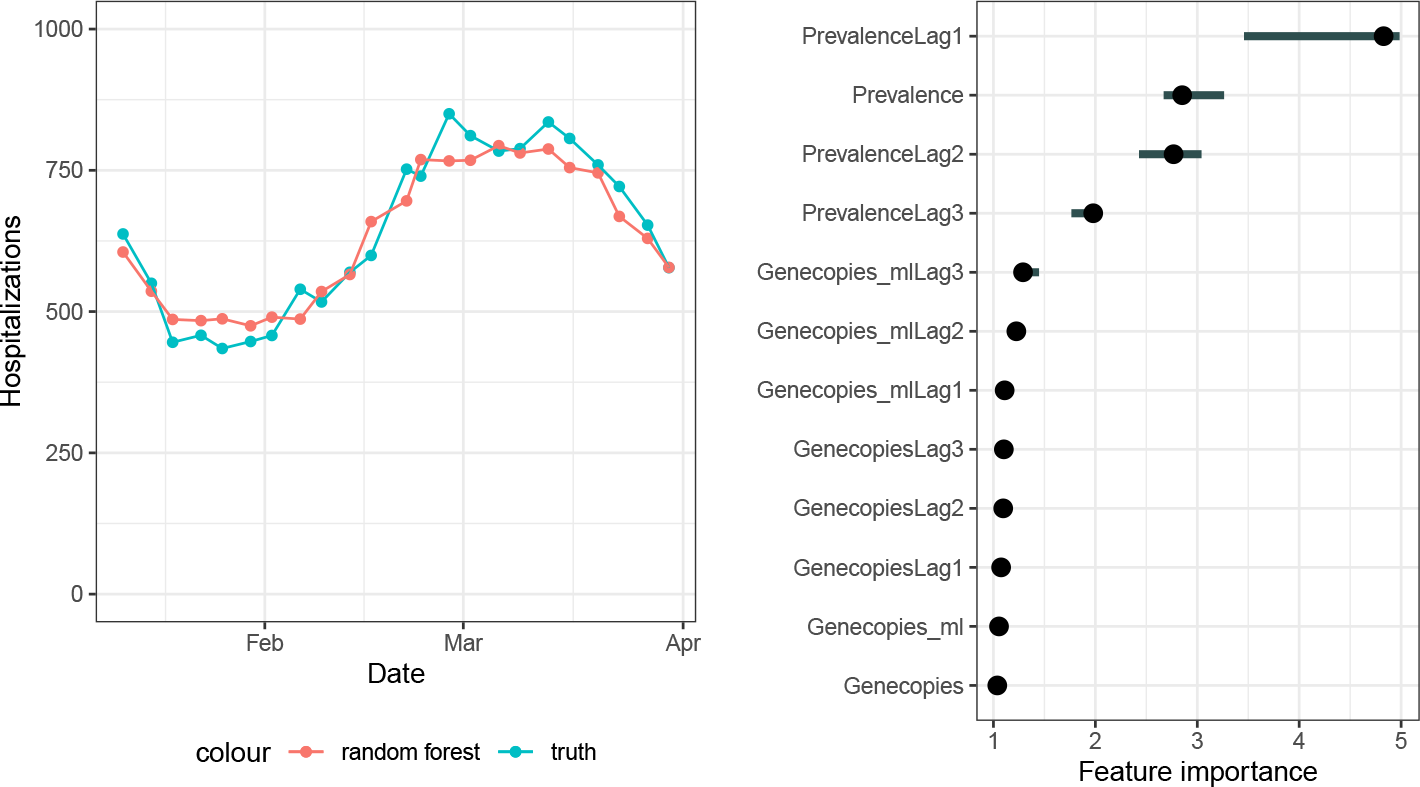
Hospitalization prediction and corresponding feature importance derived by a random forest. The number of hospitalizations can be predicted well. The most important predictors are the cohort study prevalences.

### 3.4 Finding 4: Wastewater data alone does not allow quantitative prediction of the cohort study prevalence

As observed in Section 3.3, the cohort study prevalence is a good predictor of the number of hospitalizations and, therefore, it seems to be a good indicator to investigate the development of the pandemic. Since it is less effort to test the wastewater than conducting an ongoing cohort study, it is of interest whether wastewater data can give a good estimation of the cohort study prevalence.

For this prediction, additional parameters as the air temperature or the water temperature could be used as well since the prediction is done on city level and these parameters cannot be aggregated on federal state level (cf. Section 2.1.1). We did the prediction for the cities of Koblenz, Kaiserslautern, and Mainz since for those, the investigated wastewater parameters are available. The prevalence was predicted for each city individually and the prevalence estimator for Rhineland-Palatinate was computed by combining the individual prevalences. As described in Section 2.2.2, random forests with leave-one-out cross-validation were fitted.

Figure 4 shows the results. The left panel compares the true prevalence with the random-forest-predicted prevalence. The cohort study prevalence cannot be estimated well from the wastewater data. In particular, the predicted prevalence appears to be quite constant and the peak in late February is not detected. In the right panel, the feature importances are depicted. The most relevant parameters are the genecopies per ml at the same measurement, the genecopies per ml two measurements before, and the absolute number of genecopies at the last measurement. Apart from the Covid-19 genecopies in wastewater, the most relevant parameter seems to be the air temperature and the flow.

**Figure 4.**
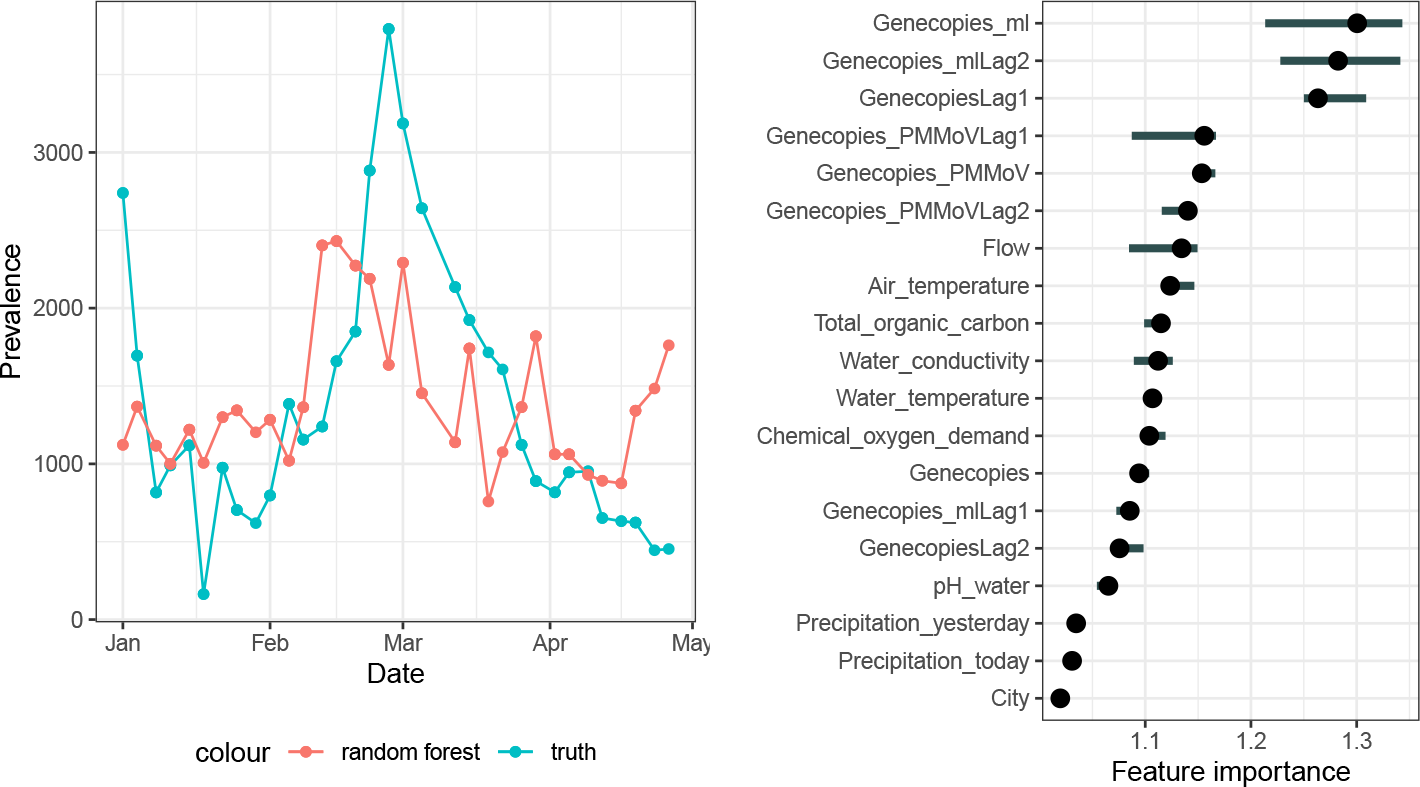
Prevalence prediction and corresponding feature importance derived by a random forest. The wastewater data do not allow a good prediction of the cohort study prevalence.

## 4 Discussion

In this paper, we described the different Covid-19 surveillance techniques that were applied in Rhineland-Palatinate, Germany. We investigated how the sources wastewater data, number of hospitalizations, and prevalence of a cohort study correlate and how they can be used to track the progress of the pandemic. It was shown that in particular the cohort study prevalence may be well suited to build a data-driven prediction model for the future number of hospitalizations.

There are some limitations that have to be noted to this observation. First, it usually takes time to take a wastewater sample and analyze the genecopies as well as to collect the results of the cohort study patients. Therefore, the data are usually not available immediately. To develop a valid prediction tool, the development of a fast data collection process would be essential. Second, the Omicron variant was the dominating variant during our entire observation period. During a pandemic, there are different variants that are dominating for a certain period of time. These variants may produce more or less genecopies in wastewater and lead to more or less hospitalizations. Consequently, a regular variant sequence analysis would be a necessary complement of an established pandemic observation tool built on wastewater. Of note, from February on, the dominated subvariant changed from BQ.1 to XBB.1. This suggests that the presented methodology is at least stable with regard to this subvariants. Third, the observed trends are mainly detectable due to the applied smoothing technique. These techniques are not valid at the boundaries of the observed time period and thus not well-suited to do extrapolation and predictions. A mathematical model that uses biological information might be a valid alternative to predict the future progress of the pandemic.

There are three possible extraction techniques for taking wastewater samples. One can extract time-proportionally, flow-proportionally, or volume-proportionally (Sandoval et al. 2018). To obtain the most representative sample, it is recommended to apply volume-or flow-proportional extraction (Wade et al. 2022). However, in our sample, all three extraction techniques were applied. To establish a regular tool for wastewater surveillance, it may be sound to apply the same extraction technique among different sewage plants and to avoid time-proportional extraction.

We aggregated the available data to the level of the federal state Rhineland-Palatinate, since the hopsitalization data was only available on this level. However, the cohort study was performed only on the five largest cities, i.e. only in urban areas and only on adults. Although these cities represent 713,000 inhabitants, i.e. 17% of the total population, and are geographically spread around the state, there may be structural differences in the infection characteristics that lead to a bias in the measured prevalence.

The wastewater treatment plants cover a size of 15,300 to 250,000 connected inhabitants, i.e., they are chosen from both rural and urban areas. In total 1,347,318 inhabitants are connected to the sewage plants, corresponding to a proportion of 33% of the total population of Rhineland-Palatinate. However, it is also not proven that these plants are perfectly representative for the federal state.

While we treat the hospitalization data as the gold standard for the burden of disease in the population, the hospitalization data only measures hospitalized people *with* a positive test and not neccessarily patients hospitalized *because of* Covid-19. It is likely, that other (respiratory) diseases such as influenza led to an overproportional rate of hospital admissions and since we measure the absolute number of positive patients, we would then see higher levels despite potentially stable disease rates.

When analyzing the information in the wastewater data by introducing other parameters than genecopies, we focused on prediction on sewage plant level and aggregated the predictions to one value for Rhineland-Palatinate afterwards (cf. Section 3.4) This was done since parameters as the air temperature or the pH value in water can hardly be summarized to one joined value among different cities. We observed a quite volatile cohort study prevalence on sewage plant level. This complicates the estimation of the prevalence and makes valid assertions on the benefit of wastewater data more difficult.

While in our data, an exact prediction of the cohort study prevalence by wastewater data without incorporating biological models was not possible, this does not imply that collecting wastewater data is senseless. Wastewater data may not be used for a quantitative prediction of the pandemic development but it may serve as an qualitative predictor. This means that an increase of the viral load in wastewater indicates a worsening of the pandemic situation. Furthermore, wastewater data can be gene-sequenced in contrast to a cohort study that measures positive Covid-19 tests. This implies that new variants or other viruses can be tested by a wastewater system as well. There already exist approaches to influenza virus Wolfe et al. (2022) and reviews on different viruses, including among other Hepatitis A viruses or noroviruses McCall et al. (2020). Since wastewater is easy and inexpensive to collect, it may develop to a more and more important tool to track the spread of different viruses in the population.

## Data Availability

The cohort study data are presented online under https://www.unimedizin-mainz.de/SentiSurv-RLP/dashboard/index.html.

The number of N1 and N2 genecopies per ml in wastewater is publicly available online under

https://lua.rlp.de/de/unsere-themen/infektionsschutz/meldedaten-coronavirus/abwassermonitoring/.

A slightly differently counted version of the number of hospitalizations than used for this paper is publicly available online under https://www.healthcare-datenplattform.de/dataset/hospitalisierung.

The German weather data are publicly available online under https://www.dwd.de/DE/leistungen/cdc/cdc_ueberblick-klimadaten.html.

https://www.unimedizin-mainz.de/SentiSurv-RLP/dashboard/index.html

https://lua.rlp.de/de/unsere-themen/infektionsschutz/meldedaten-coronavirus/abwassermonitoring/

https://www.healthcare-datenplattform.de/dataset/hospitalisierung

https://www.dwd.de/DE/leistungen/cdc/cdc_ueberblick-klimadaten.html

## Acknowledgements

We would like to thank Daniel Stich and Markus Hies for the access to all necessary data sources.

Furthermore, we would like to thank Prof. Philipp Wild and his lab for the processing of the cohort study data.

## Data availability statement

The number of N1 and N2 genecopies per ml in wastewater is publicly available online under https://lua.rlp.de/de/unsere-themen/infektionsschutz/meldedaten-coronavirus/abwassermonitoring/.

## Funding

This work has been supported by the Ministry for Science and Health of Rhineland-Palatinate.

## Notes

### Competing Interest Statement

The authors have declared no competing interest.

